# Loneliness, Social Isolation, and Effects on Cognitive Decline in Patients with Dementia: A Retrospective Cohort Study Using Natural Language Processing

**DOI:** 10.1101/2025.02.10.25321986

**Authors:** James A. C. Myers, Tom Stafford, Ivan Koychev, Robert Perneczky, Oliver Bandmann, Nemanja Vaci

## Abstract

**INTRODUCTION:** The study aimed to compare cognitive trajectories between patients with reports of social isolation and loneliness and those without.

**METHODS:** Reports of social isolation, loneliness, and Montreal Cognitive Assessment (MoCA) scores were extracted from dementia patients’ medical records using Natural Language Processing models and analysed using mixed-effects models.

**RESULTS:** Lonely patients (n = 382), compared to controls (n = 3912), showed an average MoCA score that was 0.83 points lower throughout the disease (p = 0.008). Socially isolated patients (n = 523) experienced a 0.21 MoCA points per year faster rate of cognitive decline in the six months before diagnosis (p = 0.029), but were comparable to controls before this period. This led to average MoCA scores that were 0.69 MoCA points lower at diagnosis (p = 0.011).

**DISCUSSION:** Lower cognitive levels in lonely and socially isolated patients suggest that these factors may contribute to dementia progression.

## Introduction

Social isolation (SI) and loneliness are recognised as priority public health problems,^1^ showing impact on physical and mental health,^2–7^ with effects on mortality comparable to smoking and obesity.^8^ Population attributable fraction, a measure that combines relative risk and prevalence of social isolation in a population, estimates that low social contact in older people explains up to 4·2% of the risk for dementia development.^9–12^ While much of the research has focused on SI and loneliness as risk factors for dementia^13^, these factors may also manifest as symptoms of the disease itself^14^, particularly in its early stages. In this study, we examine how SI and loneliness, when reported in the presymptomatic or symptomatic stage of dementia disease, influence the cognition of patients and the progression of the disease.

SI and loneliness are related but distinct concepts. SI is operationalised as an objective lack of social and support networks, while loneliness is seen as a negative subjective feeling resulting from the discrepancy between desired and actualised social connections and closeness to other people^1^. Participation in wider social network structures is associated with higher global cognitive function and moderates the association between cognitive functioning and amygdala volume, an indicator of neuropathological progression in Alzheimer’s disease (AD), in cognitively normal people as well as patients with Mild Cognitive Impairment (MCI) and early-stage AD^15,16^. These findings support the idea that SI influences cognitive reserve, by moderating cognitive function despite indicators of AD neuropathology.^17,18^ Feelings of loneliness have been shown to predict dementia onset^19^ and are associated with higher Amyloid Burden^7^ in cognitively normal people, as well as with higher rates of cognitive decline^20,21^ in patients with MCI, but not in participants with the diagnosis of AD^22,23^.

Despite these studies, findings for the effect of SI and loneliness on cognitive trajectories are mixed^24,25^. The main challenge in reliably estimating the impact of SI and loneliness symptoms on the cognitive function of dementia patients lies in the need for large-scale longitudinal data^26^, which is essential to capture the progressive nature of the disease.

In this study, we used Electronic Health Records (EHRs) to investigate the effect of SI and loneliness reports on the cognitive trajectories of patients with dementia diagnosis. Using textual records of patient—care provider interaction, we identify documents which discuss patients’ reports about SI and loneliness and combine them with longitudinal measures of cognitive functionality. Using data from more than 4800 patients, we estimate the cognitive trajectories of patients with reports of social isolation and loneliness and compare them to trajectories of patients without such reports, while testing cognitive changes after the first social isolation or loneliness reports. Given previous findings on the effect of SI and loneliness on the risk of dementia^9,12,20^, we expected that perceived SI and loneliness are associated with more severe cognitive decline throughout the disease.

## Methods

### Study design

The study followed a retrospective cohort design defined through the extraction of information from EHRs collected by Oxford Health NHS Foundation Trust in the United Kingdom. The data is accessible through the UK-CRIS system, maintained by Akrivia Health (https://akriviahealth.com/). The system allows access to structured information, e.g. demographic information and diagnosis code, as well as unstructured textual information, such as clinical records. These documents collect free-text information on the history of mental disorders under treatment, relevant cognitive assessments, and any other clinically relevant discussion between services and support that went on throughout the treatment.

### Cohort information

Our cohort included data from all patients with a diagnosis of Alzheimer’s or other forms of dementia (ICD codes: F00 – F00.9, F01, F02, F03, G30, but excluding F06.7 as MCI is rarely followed up in Memory Clinic). The full cohort included 34,469 patients who collectively contributed 6,388,715 medical documents from 06/03/2008 to 25/06/2022. To use the information that resides in rich clinical texts we developed Natural Language Processing (NLP) models that extracted information on cognitive health assessments in dementia and reports of loneliness and social isolation made by patients, carers, and clinical staff.

### Cognitive Outcomes

The main analysis used the Montreal Cognitive Assessment (MoCA)^27^ measure, while in supporting materials we report analysis using Mini-Mental State Examination (MMSE)^28^ scores. Both measures are widely used tools for assessing cognitive function, particularly in patients with dementia. MoCA detects mild cognitive impairments and early-stage dementia through its heavier emphasis on frontal and parietal function^29^, while MMSE, even though not as sensitive as MoCA, captures moderate to severe cognitive impairment. Both measures are frequently used in clinical practice and research^30^. MoCA scores below the cut-off point of 26 points are taken as suggestive of MCI, below 17 as suggestive of moderate impairment, and under 10 as suggestive of severe cognitive impairment. The minimum clinically important difference the smallest change in the outcome that patients would find significant, is reported to be between 0.01 and 2 points depending on the severity of the disease^31^.

### Procedure

We used structured and unstructured data from EHRs in this study. The unstructured data covered all textual records for the defined patient cohort, while structured data included information about the sex, ethnicity, and date of birth of patients, their marital and accommodation status, and ICD-10 codes for dementia and depression diagnosis (F32.0 to F34.1). To extract information about the cognition of patients, we used the previously published NLP model^32^ (for previous work on methodological considerations and description of the mental health EHR data see references^33–37^).

### NLP model for reports of SI and Loneliness

A novel NLP model was developed for the SI and loneliness reports. The model was implemented in Python and processed textual records for reports of SI and loneliness in two stages: pattern matching and classification stage. In the pattern matching stage, we used a statistical model for word processing from the Spacy library to identify words that describe SI and loneliness. This allowed us to find all documents including expressions such as “loneliness”, “social isolation”, “living alone”, etc. In the next, classification, stage, we used sentence transformer models from Huggingface’s Spacy-Setfit library to process and classify sentences with SI and loneliness mentions. Sentence transformers^38^ are types of neural network models that produce numerical representations of sentence and paragraph-level linguistic content. This vector space encodes semantic relationships, allowing us to identify and categorise semantically similar sentences. We trained sentence transformers to classify sentences with reports of SI and loneliness into four different categories: a) SI, b) loneliness, c) non-informative isolation, and d) non-informative sentences.

Reports that mention lack of social contact, living alone, and being away from family, or that mention barriers in receiving support from family, were used as an indication of social isolation^3^. Loneliness was operationalized as consisting of reports on emotional aspects of feeling lonely and suffering due to the lack of social connections^39^. The non-informative isolation category included reports of temporary and physical isolation (e.g. “isolated fall” or “isolating in the tv-room”), while the non-informative sentences category covered all incorrectly included sentences from the pattern matching stage forwarded to the sentence classification stage (see Table 1 for examples of sentences and NLP categories).

**Table 1:** Example of sentences that express the social isolation and loneliness of patients and used categorisation when training the NLP model.

| Sentence | Categorisation |
| --- | --- |
| "Is very lonely – lost husband and more recently best friend" | Loneliness |
| "Lonely and unfriended" | Loneliness |
| "Reports feeling lonely but is not trying to change this" | Loneliness |
| "Patient would wish to go out as remains isolated at home" | Social isolation |
| "Social isolation, lives in 2<sup>nd</sup> floor flat, can manage stairs" | Social isolation |
| "Due to XXX impairments and symptoms, she has gradually isolated from others" | Social isolation |
| "On the morning of the event was feeling isolated" | Non-informative isolation |
| "Alone in the tv lounge as wanting some peace" | Non-informative isolation |
| "has suffered an isolated fall" | Non-informative isolation |
| "XXX will be discharged back to your care" | Non-informative sentence |
| "Complained feeling dizzy" | Non-informative sentence |
| "Had a lovely day" | Non-informative sentence |

The full model was trained on a randomly selected subset of 11,000 medical documents from the corpus. The terms for the pattern matching stage were derived from a combination of the UCLA Loneliness Scale^40^ and linguistic phrases observed in the training set, specifically those referring to reports of SI and loneliness.

Sentences identified by the pattern matching in the training data were then used to train the classifiers.

### NLP accuracy

To evaluate the model’s performance in identifying reports of SI and loneliness, we conducted a manual annotation process using an unseen sample of 5,000 documents. The annotation was performed independently by the first and last authors. In instances of disagreement, the annotations were discussed collaboratively, and a consensus was reached to ensure consistent labelling.

The annotated data served as the ground truth for evaluating the model’s classification performance. Standard classifier performance metrics were employed, including Sensitivity, Specificity, and Balanced or F1 Accuracy. Across four different categories, the NLP model achieved average F1 accuracy of .74, reaching .83 accuracy for SI sentences (Sensitivity: .73 and Specificity: .93) and .91 accuracy for sentences reporting loneliness (Sensitivity: .88 and Specificity: .95). Full Python and R code, detailed measure of model performance, and Sensitivity analysis are reported in Supplemental materials (https://osf.io/2d5jg/).

Once trained, the NLP models were deployed on the data from the full cohort, effectively processing over 6 million medical records. To focus on the symptomatic interpretation of SI and loneliness, we only considered reports that were made 5 years before or after the initial dementia diagnosis. Our data indicates that most of these reports occur several months before and at the time of diagnosis, and that there was a substantial increase in both types of reports during the first year of the COVID-19 pandemic.

### Participants

The final data used in this study combined patients with measures of cognitive performance, as measured by MoCA scores, clinical diagnosis information, and loneliness or SI reports, consisting of 4817 patients with 9298 observations. The patient flow chart in Figure 1 outlines the procedure used to derive the sample. Our procedure identified 382 patients (851 observations) with loneliness reports and 523 patients (1185 observations) with SI reports in comparison to the 3912 patients without such mentions, which were defined as a control group in our study (see Table 2 for a split between the groups on main variables). As the retrospective cohort data were collated from EHRs, diversity, equality and inclusion could not be directly addressed during data collation.

**Figure 1.** Patient Flow Chart Illustrating Sample Derivation.

**Table 2:**
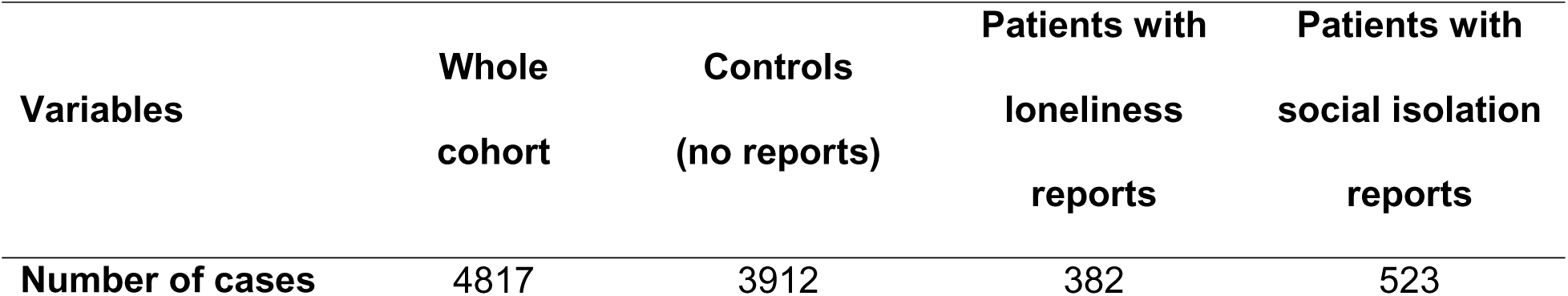
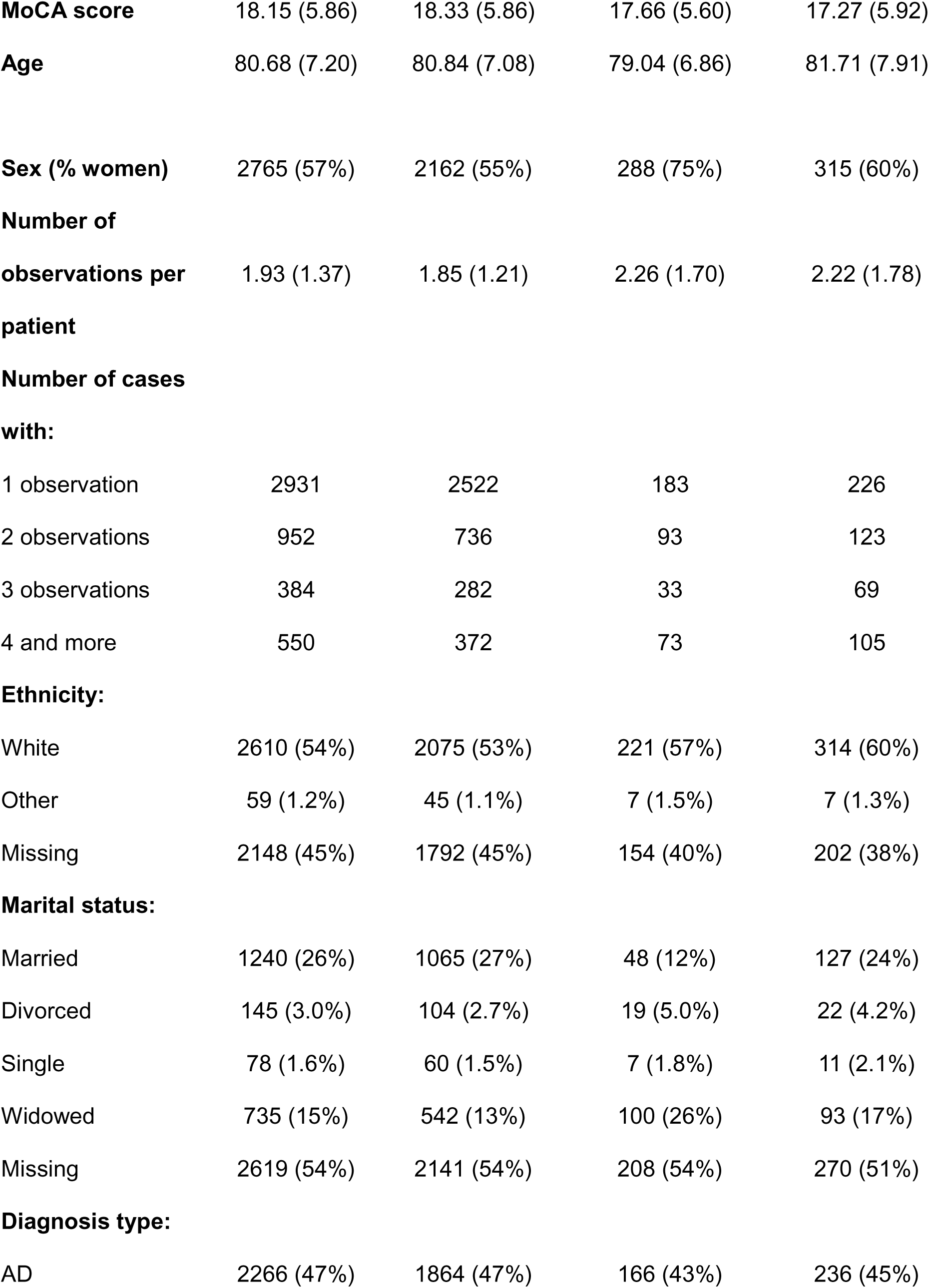
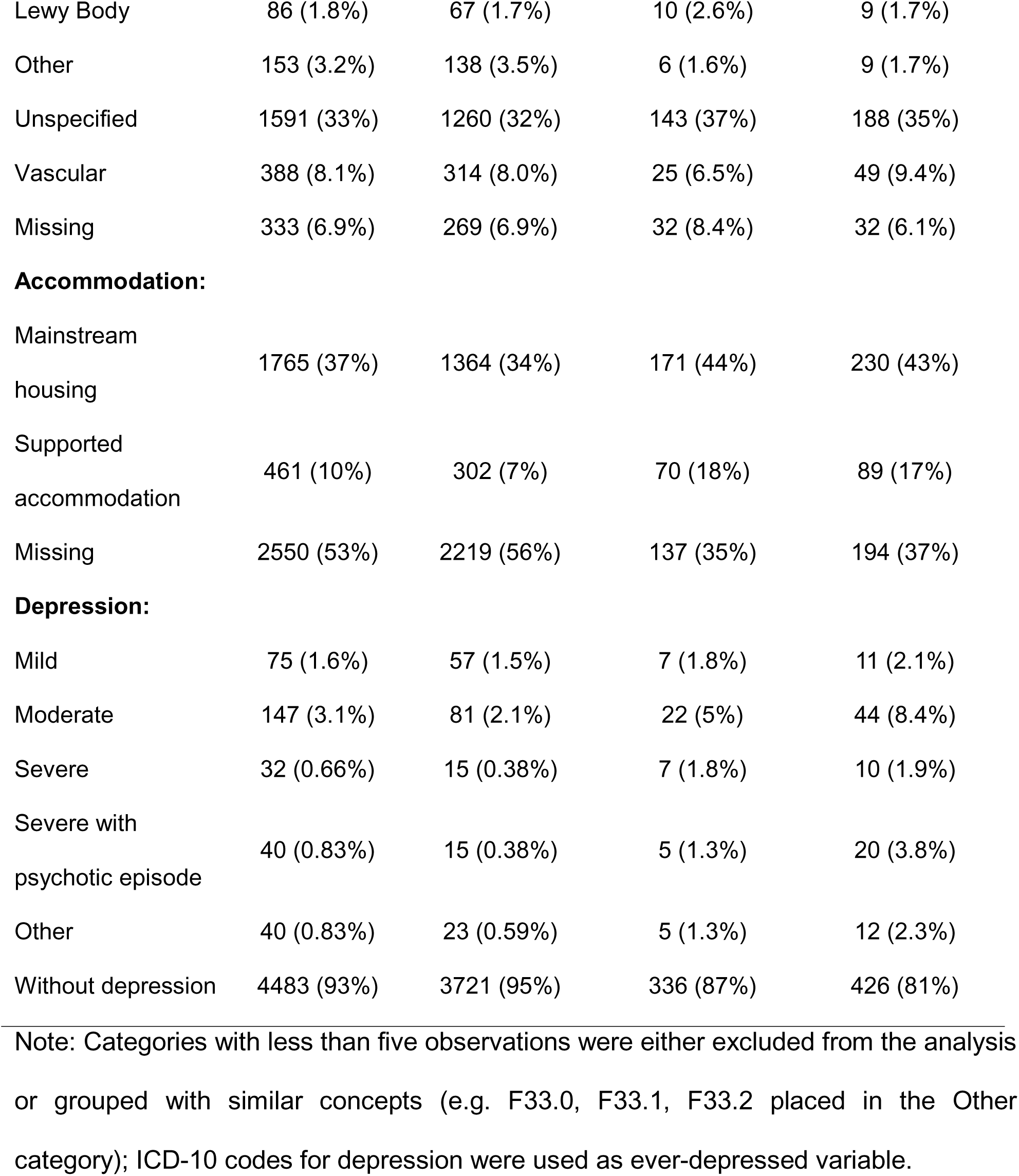
Descriptive statistics for variables of interest (Mean and SD for quantitative variables and Percentage and Number of cases for categorical variables)

| Variables | Whole cohort | Controls (no reports) | Patients with loneliness reports | Patients with social isolation reports |
| --- | --- | --- | --- | --- |
| Number of cases | 4817 | 3912 | 382 | 523 |
| **MoCA score** | 18.15 (5.86) | 18.33 (5.86) | 17.66 (5.60) | 17.27 (5.92) |
| **Age** | 80.68 (7.20) | 80.84 (7.08) | 79.04 (6.86) | 81.71 (7.91) |
| **Sex (% women)** | 2765 (57%) | 2162 (55%) | 288 (75%) | 315 (60%) |
| **Number of observations per patient** | 1.93 (1.37) | 1.85 (1.21) | 2.26 (1.70) | 2.22 (1.78) |
| **Number of cases with:** |  |  |  |  |
| 1 observation | 2931 | 2522 | 183 | 226 |
| 2 observations | 952 | 736 | 93 | 123 |
| 3 observations | 384 | 282 | 33 | 69 |
| 4 and more | 550 | 372 | 73 | 105 |
| **Ethnicity:** |  |  |  |  |
| White | 2610 (54%) | 2075 (53%) | 221 (57%) | 314 (60%) |
| Other | 59 (1.2%) | 45 (1.1%) | 7 (1.5%) | 7 (1.3%) |
| Missing | 2148 (45%) | 1792 (45%) | 154 (40%) | 202 (38%) |
| **Marital status:** |  |  |  |  |
| Married | 1240 (26%) | 1065 (27%) | 48 (12%) | 127 (24%) |
| Divorced | 145 (3.0%) | 104 (2.7%) | 19 (5.0%) | 22 (4.2%) |
| Single | 78 (1.6%) | 60 (1.5%) | 7 (1.8%) | 11 (2.1%) |
| Widowed | 735 (15%) | 542 (13%) | 100 (26%) | 93 (17%) |
| Missing | 2619 (54%) | 2141 (54%) | 208 (54%) | 270 (51%) |
| **Diagnosis type:** |  |  |  |  |
| AD | 2266 (47%) | 1864 (47%) | 166 (43%) | 236 (45%) |
| Lewy Body | 86 (1.8%) | 67 (1.7%) | 10 (2.6%) | 9 (1.7%) |
| Other | 153 (3.2%) | 138 (3.5%) | 6 (1.6%) | 9 (1.7%) |
| Unspecified | 1591 (33%) | 1260 (32%) | 143 (37%) | 188 (35%) |
| Vascular | 388 (8.1%) | 314 (8.0%) | 25 (6.5%) | 49 (9.4%) |
| Missing | 333 (6.9%) | 269 (6.9%) | 32 (8.4%) | 32 (6.1%) |
| **Accommodation:** |  |  |  |  |
| Mainstream<br>housing | 1765 (37%) | 1364 (34%) | 171 (44%) | 230 (43%) |
| Supported<br>accommodation | 461 (10%) | 302 (7%) | 70 (18%) | 89 (17%) |
| Missing | 2550 (53%) | 2219 (56%) | 137 (35%) | 194 (37%) |
| **Depression:** |  |  |  |  |
| Mild | 75 (1.6%) | 57 (1.5%) | 7 (1.8%) | 11 (2.1%) |
| Moderate | 147 (3.1%) | 81 (2.1%) | 22 (5%) | 44 (8.4%) |
| Severe | 32 (0.66%) | 15 (0.38%) | 7 (1.8%) | 10 (1.9%) |
| Severe with<br>psychotic episode | 40 (0.83%) | 15 (0.38%) | 5 (1.3%) | 20 (3.8%) |
| Other | 40 (0.83%) | 23 (0.59%) | 5 (1.3%) | 12 (2.3%) |
| Without depression | 4483 (93%) | 3721 (95%) | 336 (87%) | 426 (81%) |

Note: Categories with less than five observations were either excluded from the analysis or grouped with similar concepts (e.g. F33.0, F33.1, F33.2 placed in the Other category); ICD-10 codes for depression were used as ever-depressed variable.

### Statistical analysis

The data were analysed using a combination of generalised additive mixed-effect modelling (GAMMs)^41^ and linear mixed-effect modelling (LME)^42^. GAMM is a non-parametric data-driven method that estimates a non-linear relationship between predictors of interest and outcome variables. Using this model, we estimated and visualised average changes in cognitive function throughout the disease and tested how trajectories differed between patients who reported loneliness and SI in comparison to the control group of patients without such reports in their EHRs. To investigate the parametric effects of individual predictors on the slope of cognitive decline, we used LME.

The primary outcome in the analysis was cognitive function as measured by MoCA and MMSE tests, while we tested the effect of SI and loneliness, and controlled for the effects of Age, Type of Dementia, Sex, Marital Status, Accommodation Status, and whether patients had a diagnosis of Depression in their medical history. In case of missing information for marital and accommodation status, these patients were included in the analysis as Missing categories, while models without these predictors or cases with missing data are reported in the Sensitivity analysis. The random by-patient intercept effects were adjusted in all models, which allowed intercepts to vary for each patient^32^. The adjustment of the random-effect structure allowed us to model repeated measures of cognitive scores and guards against overfitting of the model.

Specifically, the random structure in GAMM and LME models enables us to weigh intra-individual changes over time and inter-individual differences, allowing us to estimate cognitive trajectories over time. Patients with multiple observations provide more information about the rate and direction of change, while those with fewer observations provide more information about the variability at the group level. In addition to modelling how SI and loneliness are associated with the cognitive ability of patients throughout their disease, we also explored the rate of cognitive change after patients’ first reports of loneliness and SI.

The sensitivity analysis included looking at reports of SI and loneliness in shorter time windows around the diagnosis date, different methods for dealing with missing observations in predictor variables, and MMSE score analysis. We also report analysis focusing on patients with both sets of reports, SI and loneliness, in their EHRs. In comparison to the other three groups, these patients are seen four times more frequently by the health and social services but have fewer measures of cognition, potentially indicating a more complex disease phenotype.

### Ethical approvals

We state that all procedures contributing to this work comply with the ethical standards of the relevant national and institutional committees on human experimentation and with the Helsinki Declaration of 1975, as revised in 2008. The study was approved by the local UK-CRIS oversight committees and the University of Sheffield Ethics application review board (ID: 045869). Individual patient consent was not required for the use of anonymised data. The R and Python code used to analyse the data and develop NLP models is reported in supporting materials.

## Results

The full cohort consisted of 4817 dementia patients with a mean age of 80.79 years, of whom 57% (n = 2765) were female and 26% (n = 1240) were married. Of these, 8% (n = 382) reported loneliness, 11% (n = 523) reported social isolation, and 81% (n = 3912) had no reports of either. Patients with loneliness were more likely to be women (75%) and widowed (26%), while those with SI were older (mean age 81.71 years). Controls had the highest percentage of married individuals (27%) and fewer cases of depression (4%). Table 2 reports full descriptive statistics for all variables of interest.

### The effect of loneliness and SI on cognitive trajectories

The results show a significant difference in average MoCA scores between patients with loneliness and SI reports in their EHRs and those without such reports. In the case of patients with loneliness reports, MoCA scores (see Figure 2A) were lower throughout the disease, as illustrated by Figure 2C. When estimated using LME, we see that the scores of patients who reported being lonely are lower by 0.83 MoCA points at the time of dementia diagnosis (see Table 3).

**Figure 2:** The effect of loneliness and social isolation on the nonlinear changes in cognition as measured by MoCA. A. Cognitive trajectories of the Control group (full green line) and patients with Loneliness reports (purple dashed line). B. Cognitive trajectories of the Control group (full green line) and patients with social isolation reports (purple dashed line). C. Differences in the cognitive functionality between the two groups (Average difference in MoCA scores between the control group and loneliness group) where disease periods estimated as statistically different are highlighted by the red line. D. Differences in the cognitive functionality between two groups (Average difference in MoCA scores between the control group and social isolation group) where disease periods estimated as statistically different are highlighted by the red line.

**Table 3:** Linear mixed-effect model coefficients.

|  | Estimate | Standard Error | df | t value | p-value |
| --- | --- | --- | --- | --- | --- |
| *Main Exposures* |  |  |  |  |  |
| **Intercept <sup>a</sup>** | **26.41** | **1.02** | **4463** | **25.78** | **<0.001** |
| **Diagnosis duration <sup>b</sup>** | **-0.38** | **0.04** | **7991** | **-8.71** | **<0.001** |
| **Social isolation** | **-0.69** | **0.27** | **4438** | **-2.53** | **0.011** |
| **Loneliness** | **-0.83** | **0.31** | **4381** | **-2.64** | **0.008** |
| **Diagnosis duration x Social isolation** | **-0.21** | **0.10** | **7062** | **-2.18** | **0.029** |
| Diagnosis duration x loneliness | 0.01 | 0.11 | 7495 | 0.14 | 0.885 |
| *Demographic Factors* |  |  |  |  |  |
| **Male** | **1.07** | **0.17** | **4377** | **6.15** | **0.001** |
| Accommodation Status |  |  |  |  |  |
| **Supported** | **-1.65** | **0.30** | **4213** | **5.53** | **<0.001** |
| Other | -1.40 | 0.88 | 4269 | 1.59 | 0.110 |
| **Missing** | **0.67** | **0.18** | **4391** | **3.64** | **<0.001** |
| *Clinical Factors* |  |  |  |  |  |
| **Depressed <sup>c</sup>** | **2.10** | **0.33** | **4214** | **6.28** | **<0.001** |
| Diagnosis Type |  |  |  |  |  |
| Lewy Body | -0.07 | 0.60 | 4190 | -0.12 | 0.903 |
| Other <sup>d</sup> | -0.65 | 0.46 | 4334 | -0.14 | 0.158 |
| Unspecified | 0.15 | 0.18 | 4422 | 0.85 | 0.390 |
| **Vascular** | **-1.47** | **0.30** | **4440** | **-4.82** | **<0.001** |
| **Missing** | **-1.20** | **0.49** | **4445** | **-2.43** | **0.016** |

Note: <sup>a</sup> Reference group has no complaints of SI or loneliness, is female, living in mainstream housing, not depressed, with a diagnosis of AD; <sup>b</sup> Diagnosis duration indicates the change in estimate score per every year of disease progression; <sup>c</sup> depressed variable represents patients that were ever depressed based on ICD10 codes <sup>d</sup> Type of dementia labelled Other consisted of cases with Fronto-temporal dementia, mixed Alzheimer's and Vascular dementia, and primary psychiatric disorders, while Other accommodation included clinical, no fixed abode and incarcerated.

Patients with SI reports have comparable MoCA scores to control patients before being diagnosed. Approximately six months before diagnosis, the slope of MoCA changes for SI patients becomes more severe and they start declining faster than control patients (See Figure 2B and 2D). This pattern is illustrated in the parametric model where the reference group of patients with no loneliness or SI reports show an average decline of –0.38 MoCA points per year (p < 0.001), whereas the significant interaction between SI reports and slope of diagnosis duration shows SI patients decline faster by a further –0.21 MoCA points per year relative to controls (p = 0.029). The interaction between loneliness reports and diagnosis duration was not significant, suggesting rates of decline were similar to that of patients with no SI or loneliness reports (see Table 3).

### Cognitive change after first report of SI or loneliness

The change in MoCA scores after the first SI and loneliness report behaves differently between the two groups of patients. In the case of patients reporting loneliness, the cognitive function continues to decline at the same rate as before the report of loneliness (see Figure 3A). The cognitive function of patients experiencing SI improves on average after the first mention of SI (See Figure 3B). Looking at differences in MoCA scores before and after the reports, results show that MoCA scores are on average higher after the report in the SI group (Before = 17.32 versus After = 18.08, t = 2.11, df = 1048.6, p = 0.034), but not in the loneliness group (Before = 17.36 versus After = 17.08, t = –0.67, df = 770.62, p = 0.49). However, when we identify patients with at least one MoCA measure before and after the first report and calculate repeated measure t-tests, we see that both groups of patients decline in their MoCA scores (Loneliness reports: Before = 17.58 versus After = 16.62, t = 2.31, df = 85, p = 0.023 and SI reports: Before = 18.03 versus After = 16.87, df = 115, t = 3.35, p = 0.001).

**Figure 3:** The nonlinear cognitive change, as measured by MoCA, before and after the first report for loneliness and social isolation group. A. Cognitive trajectories of patients with report(s) of loneliness. B. Cognitive trajectories of patients with report(s) of social isolation.

## Discussion

The effect of SI on the risk of dementia development is well established^9,15^, but the status of SI and loneliness when presented in presymptomatic and symptomatic stages of the disease and their effect on the progression of the disease is less explored.

We examined the effect of reported SI and loneliness on the rate of cognitive change of patients with dementia using EHRs from a UK NHS mental health trust. We showed that patients with evidence of loneliness have worse cognition throughout their disease course. Patients whose EHRs mention SI have comparable cognition, before diagnosis, to patients without mentions of such symptoms. However, six months before being diagnosed, the cognitive ability of socially isolated patients starts declining at a higher rate, resulting in worse cognition at the point of dementia diagnosis and later in the disease course.

### Mechanisms of SI and loneliness

The conceptualisation of the two concepts, SI and loneliness, likely underlies the estimated differences between the three groups of patients in our study^1^. SI represents more of a physical barrier to receiving social support and/or maintaining social connections, while loneliness reflects emotional aspects of this feeling. We found that socially isolated patients have stronger rates of cognitive deterioration several months before a dementia diagnosis while being comparable with controls in the preceding period. In the context of a life-changing and stigmatising diagnosis such as dementia, the lack of social contact prevents patients from receiving needed support, and their cognition declines at higher rates^43^. When reported for the first time, we show that average values of MoCA scores increase for this subgroup of patients. This improvement may reflect positive action taken by healthcare and/or social services in response to the report. In contrast, patients who experience loneliness have worse cognition throughout the disease course. Such results indicate that loneliness might be an intermediary for depressive symptomatology or is caused by common origins^44–46^. This interpretation is also supported by the demographic differences split by group, where we see that patients with reports of SI and loneliness also have higher rates of ICD-10 codes for depression. These patients also do not observe any improvement in their cognition after the first report of loneliness, which might be expected given that improvement in feeling of loneliness requires a change in the availability of social networks, psychological well-being, life satisfaction, activities, and other psychiatric symptoms^47^.

### Strengths and limitations

Our study illustrates the potential of medical records from mental health institutions to provide evidence-based results on the effect of symptoms in dementia diseases^9,48^. Using large data and advanced statistical modelling, our study shows that SI can be seen as a disease progression factor. Not only is this a novel finding in the domain, contrasting with some previous studies which showed limited effects or their complete absence^20,22,23^ but we show differing effects of SI and loneliness on the cognitive trajectories of patients with dementia.

There are several limitations when using large observational datasets^49^. We cannot allocate patients’ membership in the group and can only control for a limited number of factors that could have moderating effects on SI and loneliness, such as depression. Lack of control over the allocation of patients and barriers when accessing healthcare may have led to inadequacies regarding diversity, equality and inclusion, which may reduce the wider generalisability of findings. Similarly, patients with SI/loneliness reports recorded considerably before diagnosis may be a sign of additional health care needs and could additionally limit the generalisability of findings. The correlational nature of the data limits the causal interpretation of our findings, and even though we see improvement in cognition after the first report of SI, we cannot ascertain what change to patients’ social circumstances followed.

There are multiple sources of support that patients can receive after SI is identified, such as closer family connections, social services provision of care or change to their living conditions (e.g. admittance to residential care). Our interpretation of SI relies on a subjective perception from the patient, carer or clinician, rather than an objective measure e.g. of frequency of social contact. Automated and trained NLP model architectures are probabilistic, and even though they achieve high levels of accuracy, they introduce an additional layer of noise to the later data analysis^50^.

### Clinical implications

While acknowledging limitations, we show that SI and loneliness could be seen as a disease progression factor for dementia patients, given their effect on cognitive trajectories, even before a formal diagnosis. This means patients experiencing SI or loneliness might benefit from closer monitoring of their cognitive health. While cognitive decline is expected in dementia, the rate of decline can have significant impacts on both patient care and quality of life. By recognising SI and loneliness as potential factors influencing the speed of decline, these findings could have direct implications for clinical practice, such as identifying potential avenues for intervention, informing treatment strategies, and strengthening rationale for social prescribing. We hope that these effects could steer the debate concerning modifiable symptoms, as part of a holistic assessment, that could be used to support the care of patients and outline a research approach that could provide us with more evidence-based studies of modifiable disease progression factors.

## Data Availability

All patient data used in the present study are accessible through the UK-CRIS system. The source data for this work is owned by Oxford Health NHS Foundation Trust using anonymised patient records via CRIS Powered by Akrivia Health. The data cannot be made publicly available but can be accessed with permissions from Oxford Health NHS Foundation Trust for UK NHS staff and UK academics within a secure firewall, in the same manner as the authors. The R and Python code used to analyse the data and develop NLP models is reported in supporting materials.

## Consent Statement

Informed consent was not required for the study as we utilised anonymised patients’ electronic health records. The data were accessed and analysed in accordance with applicable legal and ethical guidelines to ensure the privacy and confidentiality of all subjects.

## Contributions

JM and NV were granted access to the Electronic Health Records. NV developed Natural Language Processing models, and JM supported their training and evaluation. JM cleaned and processed extracted data, JM and NV jointly analysed data, and All authors worked on the interpretation of the results. IK, RP, and OB supervised the clinical interpretation of the findings, while TS supervised analytical procedures. JM and NV wrote the manuscript, and all authors revised and reviewed the manuscript. NV, as the corresponding author, had final responsibility for the decision to submit the manuscript.

## Data Sharing

The source data for this work is owned by Oxford Health NHS Foundation Trust using anonymised patient records via CRIS Powered by Akrivia Health. The data cannot be made publicly available but can be accessed with permissions from Oxford Health NHS Foundation Trust for UK NHS staff and UK academics within a secure firewall, in the same manner as the authors.

## Acknowledgements

We acknowledge the work and support of the Oxford Research Informatics Team, Tanya Smith (Head of Research Informatics), Adam Pill and Suzanne Fisher (CRIS Academic Support and Information Analyst), Suzanne Fisher (Research Informatics Systems Analysts) and Lulu Kane (Research Informatics Administrator). Matthew Bladen’s contribution to preparing the article for publication is greatly appreciated.

The interpretation and conclusions contained in this report are those of the authors alone. The views expressed are those of the authors and not necessarily those of the NHS, the National Institute for Health Research or the Department of Health.

